# Suitability of anthrax (*Bacillus anthracis*) in the Black Sea basin through the scope of distribution modelling

**DOI:** 10.1101/2024.04.25.24306404

**Authors:** Margarida Arede, Alberto Allepuz, Daniel Beltran-Alcrudo, Jordi Casal, Daniel Romero-Alvarez

## Abstract

The Black Sea basin has a strategic geographical location bridging Asia and Europe and depends on traditional livestock practices. Anthrax, a zoonotic bacterial disease caused by *Bacillus anthracis*, poses a significant global threat impacting public health, food security, pastoralist communities, and national economies. The disease is endemic or sporadic in the Black Sea basin, however, the study of its distribution has seldom been addressed, despite its burden and the presence of historical *B. anthracis* burial sites in the region. The viability of *B. anthracis* in a particular region is going to be influenced by multiple environmental factors, such as soil composition, climate, vegetation, and host abundance. To characterize the potential distribution of *B. anthracis* in the Black Sea basin, and therefore, the potential for anthrax outbreaks, we applied an ecological niche modelling framework using the Maxent algorithm, analyzing multiple variable combinations, and proposing a novel approach for interpreting in-risk anthrax areas. Our findings underscored the importance of host abundance to the anthrax dynamics in the region. We identified anthrax-suitable areas spanning central and eastern Türkiye, Armenia, southern Georgia, southern Russia, Bulgaria, southern and eastern Romania, Hungary, Moldova, and southern Ukraine, which align with findings from previous global and regional studies on the potential suitability of anthrax. The insights gained from our research might facilitate the development of targeted interventions and policies to mitigate the spread of this disease in pastoralist communities in the Black Sea basin.

## Introduction

Anthrax, a zoonotic bacterial disease, is caused by *Bacillus anthracis*, a spore-forming, Gram-positive, and rod-shaped bacterium [1]. While wild and domestic ungulates are the primary hosts of *B. anthracis*, it can also affect other mammals, including humans [2,3]. Ruminants are typically infected through environmental exposure by ingesting the pathogen’s spores when grazing or browsing. In humans, the most common route of transmission occurs through occupational exposure to infected animal carcasses or animal products [1].

Anthrax is present in all continents, causing high yearly mortality in domestic livestock and wild animals, along with high morbidity in humans. As a result, this disease threatens worldwide public health, food security, the livelihoods of pastoralist communities, and national economies [1]. *B. anthracis* is endemic in areas of Sub-Saharan Africa, central and southwestern Asia, Central and South America, and limited regions within the United States (US). In Europe, the disease is sporadic in animals, with a higher prevalence in southern Europe, and linked to historical foci in northern areas [2]. Across the Black Sea basin, as of 2023, anthrax remained endemic in Türkiye, Azerbaijan, Georgia, and Moldova, and it was reported sporadically in Bulgaria, Romania, Ukraine, Belarus [4], and the Russian Federation [5]. Even in endemic countries, surveillance systems for anthrax are limited, contributing to underreporting and gaps in understanding its geographic extent [6]. More importantly, organic matter, calcium richness, and a neutral to alkaline pH, characteristic of black steppe soils found in central Europe, are favourable for the viability of *B. anthracis* spores in the environment [7,8]. As the environmental availability of spores is a hallmark of *B. anthracis* exposure to hosts, characterizing its ecological niche has been proposed as a way to understand its distribution [9]. The concept of the ecological niche was first introduced by Grinnell [10] as a “limited range of ecological variables that could maintain a population without immigration” exclusive to a single species. This concept was later developed by Hutchinson [11] as a quantifiable ecological area that determines species fitness and survivorship [12]. By studying the *B. anthracis* ecological niche, we aim to describe the environmental patterns that support anthrax spores’ survival which eventually leads to hosts’ exposure in the Black Sea basin [7,13].

Traditional ecological niche modelling (ENM) relies on abiotic predictors (e.g., climate) to characterize a species distribution and considers biotic interactions (e.g., host dynamics) to have negligent effects in modelling, a hypothesis called the Eltonian noise effect [14]. However, there is growing evidence that its inclusion can be crucial to describe broad-scale species distributions, especially when modelling a disease system [15]. In this study, we explored ecological niche modelling approaches based on various combinations of predictor variables, incorporating only abiotic (climate, soil, and vegetation) or introducing a biotic predictor (ruminant abundance) to assess whether the inclusion of ruminant abundance improved model performance. Additionally, we proposed a novel approach to visualize and interpret Maxent algorithm outputs by leveraging uncertainty levels to further refine the output. This allows us to suggest high-risk areas of potential *B. anthracis* outbreaks in the Black Sea basin with higher accuracy, which can guide decision-makers to prioritize awareness campaigns, surveillance, and control activities.

## Methods

This study explores the potential suitability of anthrax in the Black Sea basin through distribution modelling, using anthrax occurrences in domestic animals, from nine countries of the region, namely: Armenia, Azerbaijan, Belarus, Bulgaria, Georgia, Moldova, Romania, Türkiye, and Ukraine.

### Occurrence data and geoprocessing

We curated a database of *B. anthracis* confirmed georeferenced occurrences causing disease in domestic animal species (i.e., cattle, sheep, goats, swine, and equine) that have been reported in the participating countries between 2006 and 2021 (hereafter anthrax occurrences). The data were procured internally by FAO, sourced directly by national experts, or available online. The consolidated database included international repositories, such as EMPRES-i and the World Animal Health Information System (WAHIS), regional sources, as the Animal Disease Information System (ADIS), and national databases from Moldova and Türkiye. Finally, it includes anthrax occurrences from Deka *et al.* [16] (S1 File and S1 File Table 1).

**Table 1.** Parameters of ecological niche models categorized by principal component analysis (PCA) approach. The best model for each approach was selected using a three-step selection framework (i.e., pROC, omission rates, and AICc). AICc: Akaike information criterion corrected for sample; dem variable: demographic variable; Features: L=linear, LQ=linear+quadratic, LQP=linear+quadratic+product; PCA: principal component analysis; pROC: partial area under the Receiver Operating Characteristic; OR: omission rate; RM: regularization multiplier.

| Approach | Selected features | Selected RM | No. Predicted pixels | pROC significance | OR-5% | AICc | No. of parameters |
| --- | --- | --- | --- | --- | --- | --- | --- |
| **Approach 1:**<br>All variables<br>PCA | LQP | 0.1 | 34,917 | <0.05 | 0.0294 | 4,732.26 | 20 |
| **Approach 2:**<br>PCA by domain<br>(env only) | LQP | 0.5 | 25,895 | <0.05 | 0.0441 | 4,634.63 | 41 |
| **Approach 3:**<br>PCA by<br>domain + dem.<br>variable | LQ | 0.5 | 34,323 | <0.05 | 0.0147 | 4,715.51 | 18 |

Anthrax occurrence locations were processed in R Statistical Software (v4.2.1) [17]. We started by removing duplicates based on location and excluding records with a level of precision of less than three decimal degrees of latitude or longitude. Finally, to avoid overfitting due to spatial autocorrelation and sampling bias [16,18], we applied a spatial thinning method of 30 km [19], using the R package *SpThin* [20]. The resulting thinned occurrences were used to develop ENMs, the final dataset comprised 226 occurrences (Fig 1).

**Fig 1:**
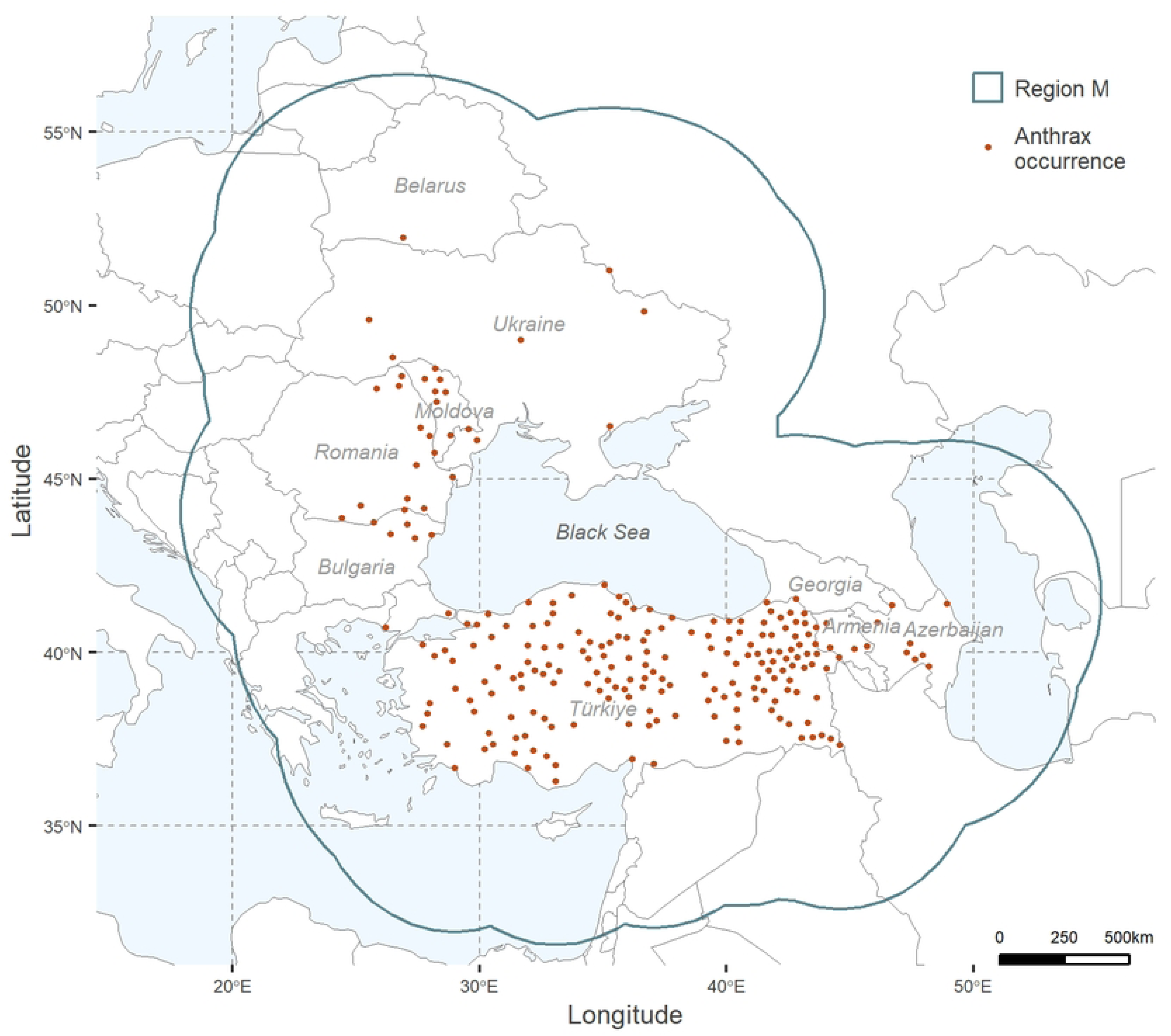
Anthrax georeferenced occurrences and calibration area (region M). *Bacillus anthracis* confirmed georeferenced occurrences (in dark orange) considered for the calculation of parameter M (outlined in teal). Maps were developed using R Statistical Software (v4.2.1) [17].

### Calibration area

The calibration region, or parameter M, is the area used to calibrate the model. The correct delimitation of M is critical as it may impact any step of an ENM, from its parameterization, validation, and model comparison [21], to the modelling outputs [22,23]. M should combine a spatial extent and environmental diversity that has been accessible to the studied species [24] during a time period that is relevant to the study [16,21]. Here, we defined M by a buffer surrounding the occurrences which distance was calculated as the mean of the distances from each occurrence to the geographic centroid [25] (Fig 1).

### Variable selection

*B. anthracis* environmental and demographic predictors were identified based on previous literature studying anthrax spatial distribution [6,19,26]. We selected four environmental categories relating to climate (i.e., temperature and moisture), soil, and vegetation, plus one demographic variable. We included 15 bioclimatic variables for temperature and moisture extracted from the MERRAclim dataset [27] at a 5 arc-minute resolution for the period 2000 to 2010, which partially matched the timeframe of our occurrences. In this study, we excluded the variables describing interactions between temperature and moisture—BIO8, BIO9, BIO18 and BIO19—due to known modelling artefacts [28]. MERRAclim is a high-resolution global repository of satellite-based bioclimatic variables, offering advantages over other commonly used climate data sources for ENM, specifically, MERRAclim shows less uncertainty in interpolated values when compared with WorldClim [27].

We selected four soil-related layers—pH, cation exchange capacity, carbon content, and nitrogen—extracted from the Global Soil Information Facilities, SoilGrids, database [29], available at https://soilgrids.org/, at a 0-5cm depth and 250m resolution. SoilGrids is a repository for chemical and physical soil properties, based on a global compilation of soil profile data sets and environmental layers. It is the result of contributions from various national and international agencies and is developed by the International Soil Reference and Information Centre (ISRIC)—World Soil Information [29,30].

As a measure of vegetation greenness, we used the Enhanced Vegetation Index (EVI) [31]. EVI’s version 6.1 was obtained through the 16-day composite images from the MOD13Q1 product at 250 m resolution [31] captured by the Moderate Resolution Imaging Spectroradiometer (MODIS) sensor, located in NASA’s TERRA satellite [32]. We processed satellite images to obtain the median from a composite of satellite images from 2005 to 2021 via Google Earth Engine [33]. EVI offers advantages over the Normalized Difference Vegetation Index (NDVI) in correcting atmospheric conditions and background noise [31].

Finally, we included a demographic variable representing ruminant abundance, resulting from the sum of three raster layers for cattle, sheep, and goats abundance sourced from the Gridded Livestock World Distribution (GLW4) and adjusted to FAOSTAT 2015 country totals at 1km resolution [34–37]. All variables were resampled to 1km resolution using the *resample* function and bilinear method in R. Further details on anthrax environmental predictors and data sources are detailed in S1 File Table 2.

To reduce high dimensionality and variable autocorrelation, we used a principal component analysis (PCA) [9,38]. We used different sets of PCAs to determine three ENM approaches. For the first approach, we calculated principal components (PCs) for the entire set of 20 environmental variables. The two other approaches comprised PCs for each environmental domain (i.e., temperature, moisture, soil, and vegetation). The third approach treated environmental domains as in the second approach, also including the ruminant abundance variable. For each of these approaches, we used the PCs retaining at least 90% of the variation in the original data [39]. PCAs were developed using the *‘kuenm_rpca’* [40] function from *kuenm* package in R [40].

### Ecological Niche Modelling

Maximum Entropy algorithm (MaxEnt version 3.4.4) [41] was implemented to define the ENMs. For this purpose, we applied the package *kuenm* [40] (https://github.com/marlonecobos/kuenm) in R Statistical Software (v4.2.1) [17] to calibrate MaxEnt ENMs and select optimal parameters for each of the three combinations of PCs as described earlier. We investigated different parameters, including combinations of MaxEnt feature classes (i.e., response types: linear, linear+quadratic, linear+quadratic+product), and five regularization multipliers (i.e., 0.1, 0.5, 1, 1.5, and 2).

#### Model evaluation

We partitioned anthrax occurrences randomly: 70% of occurrences for model training (calibration), and 30% of occurrences for model testing (evaluation) [42,43]. Models were primarily evaluated and selected via the *kuenm* package [40] following a three-step approach. First, models were assessed for statistical significance (*p*-value<0.05) based on the partial area under the curve of the Receiver Operating Characteristic (*pROC*). Then, those models with a lower omission rate (OR, threshold=5%), [44], were selected. Lastly, the resulting models were further narrowed down using the Akaike information criterion corrected for sample size (*AICc*) [45] to ensure low model complexity and good fit to the underlying data.

#### Final Model

Final models were generated with the function *‘kuenm_mod’* from *kuenm* [40]. For the three modelling approaches, we specified the output format as logistic, with a continuous scale from 0 (non-suitable) to 1 (suitable). Additionally, we used 50 bootstrap replicates to calculate the median and assess model uncertainty, i.e., the difference between the rasters with maximum and minimum values. Final model outputs were categorized (i.e., suitable vs. non-suitable) considering the suitability value from the 95% of the calibration points (E=5%) as threshold for binarizing the model [46].

From the three modelling approaches, we selected the best model based on the following criteria: lowest OR, lowest number of parameters, larger predicted area, and lowest uncertainty. Finally, to interpret the final model, we overlapped the best binarized model (i.e., suitable/unsuitable) with the uncertainty raster and considered highly suitable areas to those with less than the third quartile of uncertainty values.

## Results

A total of 1182 raw anthrax outbreak occurrences in domestic livestock, spanning from 2006 to 2021, were collated from various sources and used in the current study (S1 File. Table 1). Cattle, sheep, and goats outbreaks accounted for 80.7%, 14%, and 4% respectively, representing the majority of studied outbreaks (98.7%). The remaining occurrences represented outbreaks attributed to horses and swine (1.3%). Over the studied period, the cumulative frequency of anthrax occurrences started increasing in July, peaked in September (n=193) at three times the mean for the first six months of the year (n=65), and gradually decreased until December (n=62, S2 Fig).

Each of the three explored approaches resulted in 15 candidate models, reflecting combinations of three feature classes and five regularization multiplier values. The three best-fitting models were identified through the described three-step framework (Table 1).

The model output for *B. anthracis* developed using a PCA per environmental domain plus the variable representing ruminant abundance in the studied area were selected as the best overall model (i.e., approach 3; Table 1). This model yielded a wider prediction with lower uncertainty and presented a lower OR with a lower number of parameters than the two other approaches (Table 1). To generate this ENM approach, we retained the first three PCs for temperature and soil, explaining 98.83% and 95.77% of their respective domains, the first two PCs explaining 99.44% of the moisture domain, and one PC each for EVI and ruminant abundance. Models’ median, uncertainty, and areas suitable and non-suitable for *B. anthracis* at 5% threshold are illustrated in Fig 2. Outputs for the other two approaches can be found in the S2 File Fig 1. We highlight that the temperature and soil domains had the highest contribution to the final selected model accounting for 38.2 and 32.9%, whereas similar contributions were attributed to EVI and ruminant abundance, at 10.3% and 9.9%, respectively (S2 File Table1).

**Fig 2:**
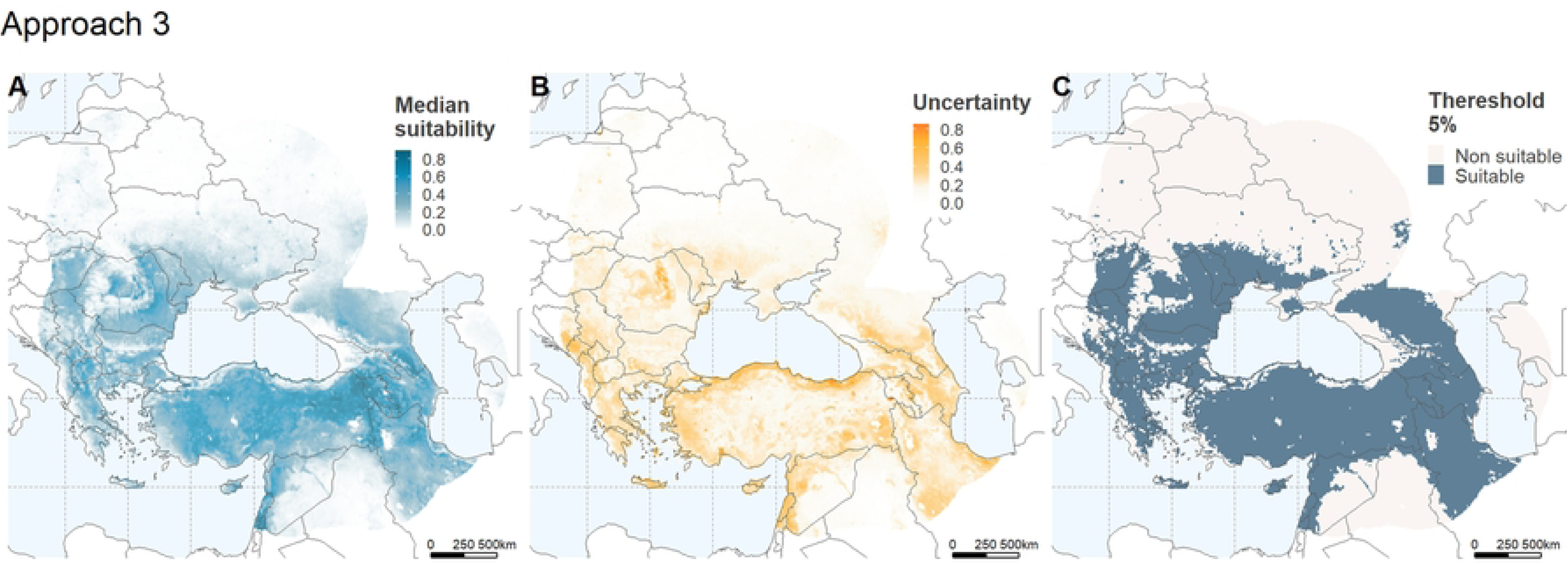
Ecological niche model outputs for *Bacillus anthracis* in the Black Sea basin. Model outputs for the selected best model for *B. anthracis* using principal components (PCs) by domain plus the demographic variable based on ruminant abundance (i.e., approach 3; Table 1). Maps depict (A) continuous suitability, (B) uncertainty, and (C) a binary map of suitability using a 5% threshold. Maps were developed using R Statistical Software (v4.2.1) [17].

We contrasted suitable areas for anthrax in the overall best model binary map with varying levels of model uncertainty. Low uncertainty was defined here as those pixels with values below the third quartile of the uncertainty range (i.e., Q3= 0.23; Fig 3A). Regions identified as highly suitable with low uncertainty (Fig 3B) span western to central Armenia, extending into the southwest of Azerbaijan; they include a limited area in the northeast of Azerbaijan and the southern border region of the Russian Federation; the interior regions of the Islamic Republic of Iran and southern Russian Federation; as well as the interior eastern, central and central-south areas of Türkiye (Fig 3B). Additionally, anthrax suitability is also observed in centre south and north Bulgaria and south and east Romania, centre east of North Macedonia, north of Serbia, southeast of Hungary, centre to south of Moldova, and the south coast of Ukraine with the Black Sea (Fig 3B).

**Fig 3.**
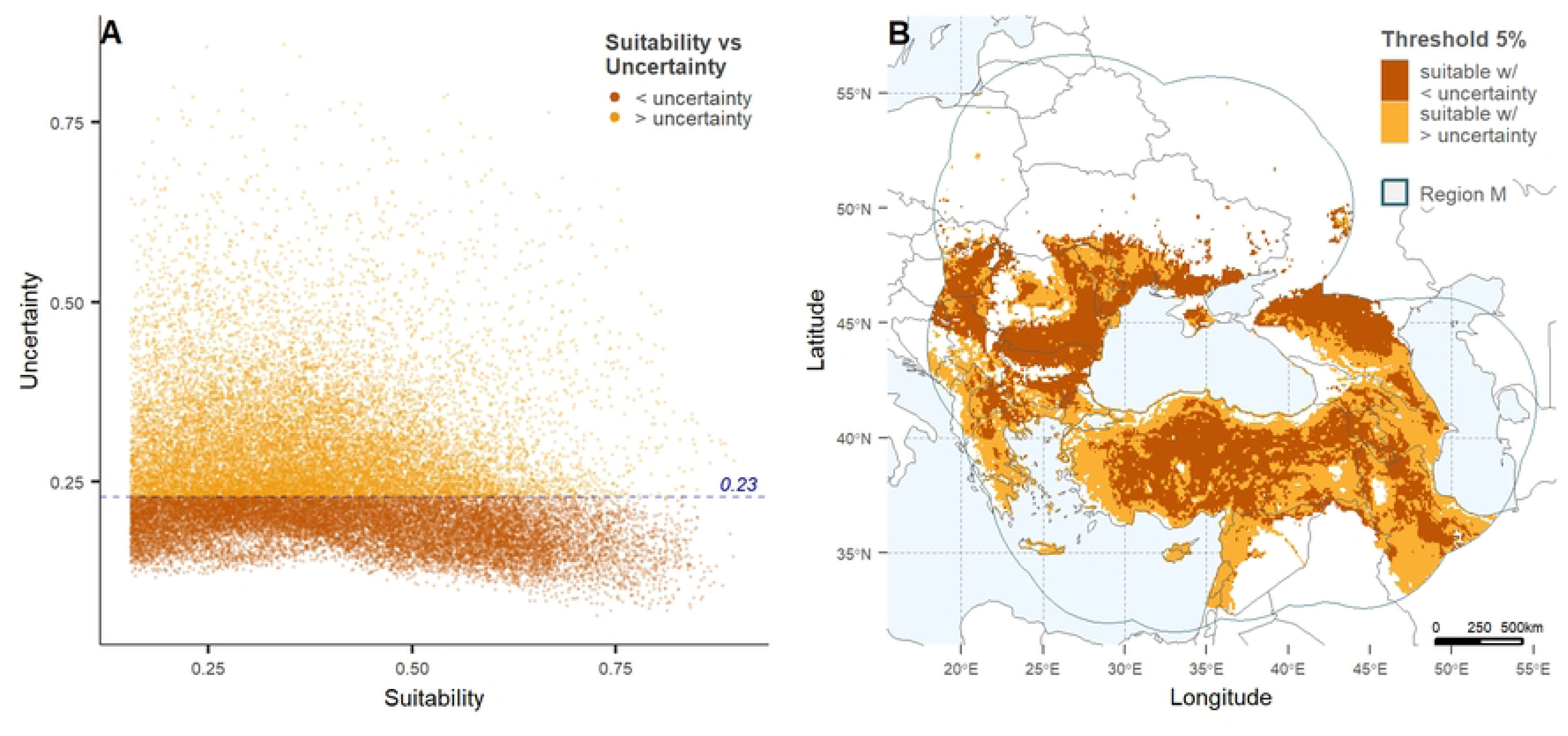
Suitability versus uncertainty regions for the best-selected model of the potential distribution of *Bacillus anthracis*. (A) Illustrates the correlation between continuous anthrax suitability and uncertainty for the best model (Table 1, Fig 2). High uncertainty was defined by a cut-off set as the third quartile across all uncertainty values (>=0.23). (B) Depicts the 5% binary output of anthrax suitability with higher (orange) and lower (ochre) uncertainty. Graph and map were developed using R Statistical Software (v4.2.1) [16].

Regions with high suitability with low uncertainty where no anthrax occurrences have been reported (Fig 1 and Fig 3B) can be found in the southern interior of the Russian Federation, the interior of the Islamic Republic of Iran, the central southern region of Bulgaria, central-east of North Macedonia, northern Serbia and centre to east of Hungary. Conversely, regions where anthrax cases have been reported, yet are depicted in our models as areas of low anthrax suitability, are primarily seen in central to northern regions of Ukraine and southern regions of Belarus. High suitability areas with high uncertainty are observed along the coast of southern Türkiye with the Black Sea, the west coastal area of Türkiye with the Mediterranean Sea, and the southern-east region of Türkiye along the border of the Republic of Iraq and the Islamic Republic of Iran.

## Discussion

Through the scope of distribution modelling, we found highly suitable regions for *B. anthracis* survival in the Black Sea basin; these areas might well benefit from investment and resource allocation for the control and prevention of anthrax outbreaks. Our model’s predictions agreed with findings from previous studies conducted at various geographical scales. Suitable areas identified for anthrax spanned from central to eastern Türkiye, Armenia, southern Georgia, the southern Russian Federation, Bulgaria, southern and eastern Romania, Hungary, Moldova, and southern Ukraine. These areas are similar to those found by recent studies exploring the ecological niche of *B. anthracis* at a global scale [6,16], as well as a study specifically focused on northern latitudes [47]. Additionally, our model found anthrax-suitable areas with low uncertainty in northeast Azerbaijan, consistent with anthrax spatial clusters observed between 2000 and 2010 [48]; and the Odesa region in Ukraine, converging with a publication reporting *B. anthracis* in environmental samples and animal anthrax cases in this area [49]. Finally, we should highlight that although our model did not include anthrax occurrences from Georgia, it accurately predicted the southeastern region of this country as suitable for anthrax, corroborating previous reports (Pers. Comm. T. Chaligava). However, it was unable to predict similar suitability in central to northern regions of Georgia, where both livestock (Pers. Comm. T. Chaligava) and human anthrax cases [50] have been documented.

There is a well-established spatio-temporal link between human and livestock anthrax cases due to the high occupational nature of anthrax in humans [1]. In this regard, our model corroborates the high incidence of human and livestock anthrax cases found in eastern provinces of Türkiye, clustering around animal trade centres and large international commercial roads [51,52] and linked with livestock trade routes between eastern and western Türkiye and from the centre Anatolia to the southern and northern parts of the country [52].

Upon comparing Maxent ENMs assessing various variable combinations, we found that the inclusion of the ruminant abundance (biotic variable)—which PC ranked fourth in the final model (S2 File Table 1)—improved model performance and was an important parameter in selecting the best overall model of anthrax suitability in this region. Livestock’s abundance has previously been explored and seen as influential in anthrax distribution studies [26,47,53–56]. These results emphasize the importance of biotic interactions for disease systems [15]; ruminants are the most susceptible hosts to *B. anthracis* and play a key role in the maintenance and transmission of anthrax [57]. It is worth noting that ruminant production is a critical livestock subsector in the majority of the studied countries [58–67]. In addition, areas found as suitable for anthrax by our model largely match rural settings where pastoralism is widely practiced [68], and livestock is the main source of subsistence for these populations [4,68]. Similarly, Carlson *et al* [6] suggested higher human anthrax risk in rural areas, and observed increased human and livestock anthrax vulnerability in rainfed systems across arid and temperate landscapes in the same region (Eurasia).

Soils and temperature had the highest contribution percentage to our model (S2 File Table 1). Chernozem or black steppe-type soils, prevalent in eastern Europe [69] and partly covering our M region, are known to create favourable conditions for anthrax sporulation [70] and have been associated with anthrax epidemics [7]. At the same time, the southern part of the M region, where the mean annual temperature is higher, was identified as suitable for anthrax by our model. This result aligns with established knowledge regarding favourable conditions for anthrax viability in areas with temperatures exceeding 15 ⁰C [3] and is further supported by results from Carlson *et al*. and Walsh *et al.* [6,47]. Furthermore, cumulative anthrax occurrences were higher between July and October. This period corresponds to high temperatures and dry conditions across the region [71], which facilitate the mechanical dispersion of anthrax spores [8]. Additionally, this period coincides with the time when ruminants graze in local pastures or migrate to summer pastures. As the grass gradually becomes shorter during this season, ruminants tend to graze closer to the soil, heightening their risk of exposure to the *B. anthracis* spores [72]. Moreover, the high temperatures during this time may also lead to ruminants’ nutritional stress and compromise their immunocompetence, making them more susceptible to the disease [73]. Such temporal pattern was previously observed in Azerbaijan [74], Türkiye [75] and Kyrgyzstan [72].

Some of the few anthrax occurrences in the northern M region were missed by our final model (Fig 3). This discrepancy may be attributed to the low mean annual temperature at these latitudes, which theoretically hinders anthrax viability [3]. However, it is worth noting that during summer months, temperatures may still enable significant sporulation of *B. anthracis* [3]. In contrast, Deka *et al.* [16] showed “very high” and “high” suitability for anthrax in parts of our northern region M, diverging from our findings. Additionally, anthrax cases in Ukraine and Belarus were reported sparingly, likely due to rigorous documentation of biothermal pits and infected burial grounds [49]. These areas are subject to strict legislation prohibiting any construction as well as agricultural and pastoral practices without prior disinfection at these sites. Furthermore, the lack of cases in these countries may be also explained by the prevalence of intensive livestock production systems where ruminants are often confined, and pastoral practices are uncommon, reducing opportunities for exposure to anthrax spores. Nevertheless, despite the current suboptimal environmental conditions for anthrax viability in this region, climate change- led extreme weather events, such as warmer temperatures, high precipitation and droughts [76] are expected to increase anthrax risk in these areas [16,47].

Besides local climate, soil characteristics, host demography, and wildlife interactions, anthrax outbreaks are associated with a range of socio-economic factors. These factors encompass food security, disease awareness, cultural and religious events, as well as access to veterinary services and healthcare. These factors are directly linked with livestock production practices, including production systems, pastoralism, seasonal movements, veterinary surveillance and control capacity, vaccination use and coverage, and the application of biosecurity measures [77]. Further research into the impact of these factors on the risk of anthrax outbreaks among livestock and humans in the region would complement the findings of the current study.

Our regional-scale map illustrating anthrax suitability complements existing studies targeting this region at broader scales [6,16,47,56]. In our study, we explicitly incorporated uncertainty measures into our final predictions, aiming to highlight and define more accurately potential anthrax-suitable. The inclusion of uncertainty in the final outputs of ENMs is seldom implemented [6,16,19,26], and we advocate for its consideration, especially in ENM studies exploring pathogens.

As an evidence-based map of anthrax distribution, the areas highlighted by our model should guide future research efforts aimed at anticipating future outbreaks. They should facilitate resource allocation to improve the cost-efficiency of surveillance and control activities, as well as disease awareness and educational campaigns promoting appropriate quarantine, carcass handling, and disposal. For the success of such preventative measures, we stress the importance of coordinated efforts between the veterinary and public health sectors at both national and international levels.

## Conclusions

Our study identified high-risk areas for anthrax across central and eastern Türkiye, Armenia, southern Georgia, southern Russia, Bulgaria, southern and eastern Romania, Hungary, Moldova, and southern Ukraine. These findings are critical for prioritizing resource allocation and implementing anthrax management interventions in the region.

Leveraging uncertainty levels and explicitly including them in our modelling approach improved the reliability of the potential suitable and non-suitable regions for anthrax identified in our final maps. We believe this approach also facilitates the interpretability of our results and enhances their utility for decision-makers and stakeholders.

The inclusion of ruminant abundance as a biotic variable in our modelling framework significantly improved model performance, highlighting the importance of host-pathogen interactions in the study region.

Overall, anthrax poses a significant threat to livestock, particularly ruminants, whose production sector is essential for the economies and subsistence of rural populations in the Black Sea region. We anticipate that the risk maps generated in this work offer comprehensive insights into anthrax distribution in this region, providing valuable guidance for targeted interventions to mitigate the impacts of this disease.

## Data Availability

The data that support the findings of this study are available from national authorities of the participating countries and Animal Disease Information System (ADIS). Restrictions apply to the availability of these data, which were used under licence for this study. Data are available with the permission of data owners.

## Acknowledgements

The authors are thankful to the relevant national authorities from the participating countries for sharing their data to carry out the project. We acknowledge Giuseppina Cinardi from FAO-NSAL for her contribution to generating GLW 4 maps for ruminant distribution. We would also like to acknowledge the United States DoD DTRA Cooperative Threat Reduction Program’s support of project HDTRA1-19-1-0037 “Global Framework for the Progressive Control of Transboundary Animal Diseases (GF- TADs)”.

## Supporting Information Captions

**S1 File. Anthrax occurrences data sources, environmental domains, and R packages used in the current study. (PDF)**

**S2 Fig. Seasonal trend of *Bacillus anthracis* georeferenced occurrences during our study period. (TIFF)**

**S2 File. Description of model outputs for non-selected models and Maxent output for selected approach. (PDF)**

